# Population attributable fraction of modifiable risk factors for dementia in the Democratic Republic of Congo: A community-based cross-sectional analysis

**DOI:** 10.64898/2026.03.23.26349098

**Authors:** Jean Ikanga, Caterina Obenauf, Megan Schwinne, Saranya Sundaram Patel, Guy Gikelekele, Emmanuel Epenge, Japhet Magolu Potshi, Topina Tomadia, Immaculee Kavugho, Francine Manyonga Sabowa, Joseph Phuati Tsangu, Francis Muena Kondo Beya, Samuel Mampunza, Lelo Mananga, Justine Bukabau, Thomas Karikari, Alden L. Gross, Alvaro Alonso

## Abstract

**Background:** Estimates from high-income countries suggest that approximately 40% of dementia cases may be attributable to modifiable risk factors across the life course. However, most evidence informing these estimates originates from high-income settings, and population-level estimates from sub-Saharan Africa remain limited. We aimed to estimate population attributable fractions (PAFs) for modifiable dementia risk factors in the Democratic Republic of the Congo (DRC).

**Methods:** We conducted a cross-sectional analysis of community-dwelling adults aged 65 years and older enrolled in the *Étude du Vieillissement Cognitif et de Démence en République Démocratique du Congo* (EVCD-RDC). Prevalence estimates of dementia and associated exposures were derived from prior epidemiological studies in this population. Odds ratios were estimated using logistic regression, and population attributable fractions were calculated by integrating exposure prevalence with effect size estimates. To account for correlations between exposures, communality weights were applied when estimating combined PAFs across risk factors.

**Findings:** Combined modifiable risk factors were estimated to account for 37.3% (95% CI 14.3–55.6) of dementia cases in this sample. Poverty had the largest weighted PAF (18.4%, 95% CI 13.3–22.8), followed by low educational attainment (11.3%, 95% CI 7.3–15.3) and depression (5.8%, 95% CI 2.8–8.6). Additional contributors included traumatic events (5.4%), war exposure (2.1%), diabetes (1.3%), and hypertension (1.1%). A hypothetical 15% proportional reduction in these risk factors was estimated to reduce dementia prevalence by 6.4% (95% CI 2.1–10.8), corresponding to approximately 10 700 cases prevented in the DRC by 2025.

**Interpretation:** Modifiable risk factors account for a substantial proportion of dementia burden in the DRC, with structural determinants such as poverty and education contributing the largest fractions. Dementia prevention strategies in low- and middle-income countries may therefore require broader public health approaches that address socioeconomic and structural determinants alongside conventional clinical risk factors.

**Funding:** National Center for Advancing Translational Sciences of the National Institutes of Health (UL1TR002378).

## INTRODUCTION

Dementia is a growing global public health challenge, with disproportionate projected increases in low- and middle-income countries (LMICs). More than 57 million people live with dementia worldwide, and nearly three-quarters of future cases are expected to occur in LMICs, with cases projected to nearly triple by 2050.^1^ Dementia already imposes healthcare, social, and economic costs exceeding US$1 trillion annually.^2^ An estimated 40% of dementia cases could be prevented by addressing modifiable risk factors across the life course.^3,4^ Most dementia research has been conducted in high-income countries (HICs), and the burden of dementia and its risk factors in sub-Saharan Africa (SSA) remains poorly characterized.^5^ This evidence gap limits effective public health strategies because policymakers lack region-specific data to guide interventions. Population attributable fraction (PAF) metrics estimate the proportion of dementia cases that could be prevented if modifiable risk factors were reduced.

PAF estimates the proportion of cases attributable to specific exposures. Despite its utility, PAF estimation for dementia has largely focused on HICs, with limited application in LMICs and SSA. The 2020 and 2024 Lancet Commissions identified 14 modifiable risk factors and estimated that up to 40% of dementia cases could be prevented or delayed through coordinated interventions.^3,6^ These risk factors span the life course and include early-life low education; midlife hearing loss, traumatic brain injury, hypertension, alcohol use, and obesity; and later-life smoking, depression, social isolation, diabetes, air pollution, and vision impairment.^7^

Although evidence is strongest in HICs, emerging data suggest dementia risk factors may be more prevalent in LMICs. In SSA, estimates of dementia risk factor prevalence and PAFs remain limited. The lack of SSA-specific PAF estimates represents a missed opportunity, as LMIC studies suggest greater potential for dementia risk reduction than in HICs.^5^ For example, low educational attainment is both highly prevalent and strongly associated with dementia risk in many SSA countries. In several LMICs, low education has been identified as a major modifiable risk factor with PAFs exceeding those reported for non-SSA LMICs.^8,9^

Structural determinants such as poverty, conflict exposure, and trauma are common in SSA but rarely included in dementia PAF analyses. Chronic exposure to deprivation and conflict-related stress may influence cognitive ageing through pathways involving vascular health and educational disruption. Some literature suggests that adversity may foster resilience, but chronic stress and trauma are also recognized contributors to neurobiological vulnerability and cognitive decline.^10,11^ Depression is another increasingly important dementia risk factor in SSA and is closely linked to poverty, social isolation, and other structural determinants.^3,7,12,13^ In Mozambique, for example, the PAF for depression is nearly three times that observed in other non-SSA LMICs.^8^

Non-communicable diseases that increase dementia risk—such as diabetes, obesity, and hypertension—are also rising globally, including in SSA.^3,14–17^ Among LMICs outside SSA, hypertension, diabetes mellitus, and obesity are established dementia risk factors with PAFs of 6.8%, 2.1%, and 5.3%, respectively.^7,8^ Tobacco and alcohol use also represent public health challenges: more than 80% of the world’s tobacco users live in LMICs, and smoking prevalence is rising or stagnating in several of these countries.^18–20^ Meanwhile, ambient and household air pollution is increasing in SSA and is increasingly recognized as a modifiable dementia risk factor.^21,22^ Although air pollution has not been studied comprehensively across LMICs, there is mounting evidence that it is a significant contributor to dementia risk in the Democratic Republic of Congo (DRC).^3,17^ Sleep disorders—reported in about 31% of older adults in the DRC—have also been linked to dementia risk, although PAF estimates for sleep-related risk in SSA are not yet available.^3,17^

The DRC is an important but underrepresented setting for dementia research in SSA. It is the second-largest country in Africa by area and among the most populous in the region.^23,24^ Although life expectancy in the DRC (61.6 years) remains below the African average (63.6 years), the older adult population is expanding and dementia prevalence is estimated at 6.2%.^4,23^ Over three decades of armed conflict and political instability have disrupted educational systems, entrenched poverty, and contributed to increasing rates of metabolic conditions such as hypertension and diabetes, high alcohol and tobacco use, sleep disorders, and elevated air pollution exposure—all of which are potential modifiable risk factors for dementia.^4,25–27^ In a recent sample of older adults in the DRC, 44% reported war-related experiences and 59% reported traumatic events, and individuals with suspected dementia reported higher levels of poverty than cognitively healthy peers.¹ These patterns highlight the need to examine poverty, war, and trauma alongside traditional cardiometabolic risk factors when estimating dementia risk.^9,28^

The DRC provides a context to examine cumulative exposures related to conflict, poverty, and cardiometabolic risk and to estimate PAFs for understudied dementia risk factors. Using a community-based cohort of older adults in Kinshasa, we hypothesized that PAFs would be highest for low education, depression, diabetes, and hypertension. We further anticipated that less-studied exposures—such as poverty, war experience, traumatic events, and air pollution—are also associated with dementia risk. By estimating PAFs for these modifiable risk factors, we aim to inform dementia prevention priorities in the DRC.

## METHODS

### Study population

Participants were drawn from the Étude du *Vieillissement Cognitif et de Démence en République Démocratique du Congo* (EVCD-RDC; Study of Cognitive Aging and Dementia in the Democratic Republic of the Congo), a prospective community-based cohort examining cognitive ageing and dementia among older adults in the DRC.^17^ We recruited and screened 1872 volunteers through community outreach in churches, senior associations, military camps, hospitals, and clinics in Kinshasa. This recruitment strategy did not constitute probability sampling. Cognitive screening used the Montreal Cognitive Assessment (MoCA),^29^ and functional status was assessed with the Identification and Intervention for Dementia in Elderly Africans (IDEA) instrument.^30^

As previously described in (see Figure 1)^17^, eligibility criteria were: (1) Congolese citizenship and residence in the DRC; (2) age ≥65 years; (3) provision of contact information for a caregiver or informant; (4) w willingness to participate longitudinally; (5) capacity to provide informed consent independently or via a representative; (6) fluency in Lingala or French; and (7) no history of major neurodevelopmental, psychiatric, or neurological disorders. Individuals with visual or auditory impairment preventing testing were excluded. Dementia was defined as cognitive impairment (>1.5 SD below cohort norms in ≥2 domains) with functional decline (IDEA <30). Biomarker confirmation was not required.

### Procedure

As previously reported,^17^ the study followed four phases:

1. Recruitment and enrolment. Community leaders distributed study announcements describing eligibility and study aims. Interested volunteers were approached by trained research staff for eligibility screening and consent.
2. Screening and evaluation. Screening instruments and neurocognitive assessments were administered by trained staff using standardized procedures.
3. Sample collection and neurological evaluation. Biological specimens (cerebrospinal fluid, blood, urine, saliva, and stool) and neurological examinations were obtained at the *Centre Médical de Kinshasa* (CMK).
4. Classification and adjudication. Participants were classified as cognitively unimpaired (CU), mild cognitive impairment (MCI), suspected Alzheimer’s disease (AD), or other dementia (OD). A multidisciplinary adjudication panel (neuropsychologist, neurologists, psychiatrist, internist) reviewed all data to determine cognitive status and etiological diagnosis.

Following establish criteria,^31^ participants without cognitive impairment and with preserved functioning (IDEA ≥30) were classified as CU. Clinical dementia was defined as impairment in at least two cognitive domains (>1.5 SD below cohort norms) with functional decline (IDEA <30). Progressive memory-predominant decline was classified as suspected AD; stepwise decline, fluctuating performance, identifiable medical causes, or behavioral change were classified as other dementia. (Figure 1).

**Figure 1.**
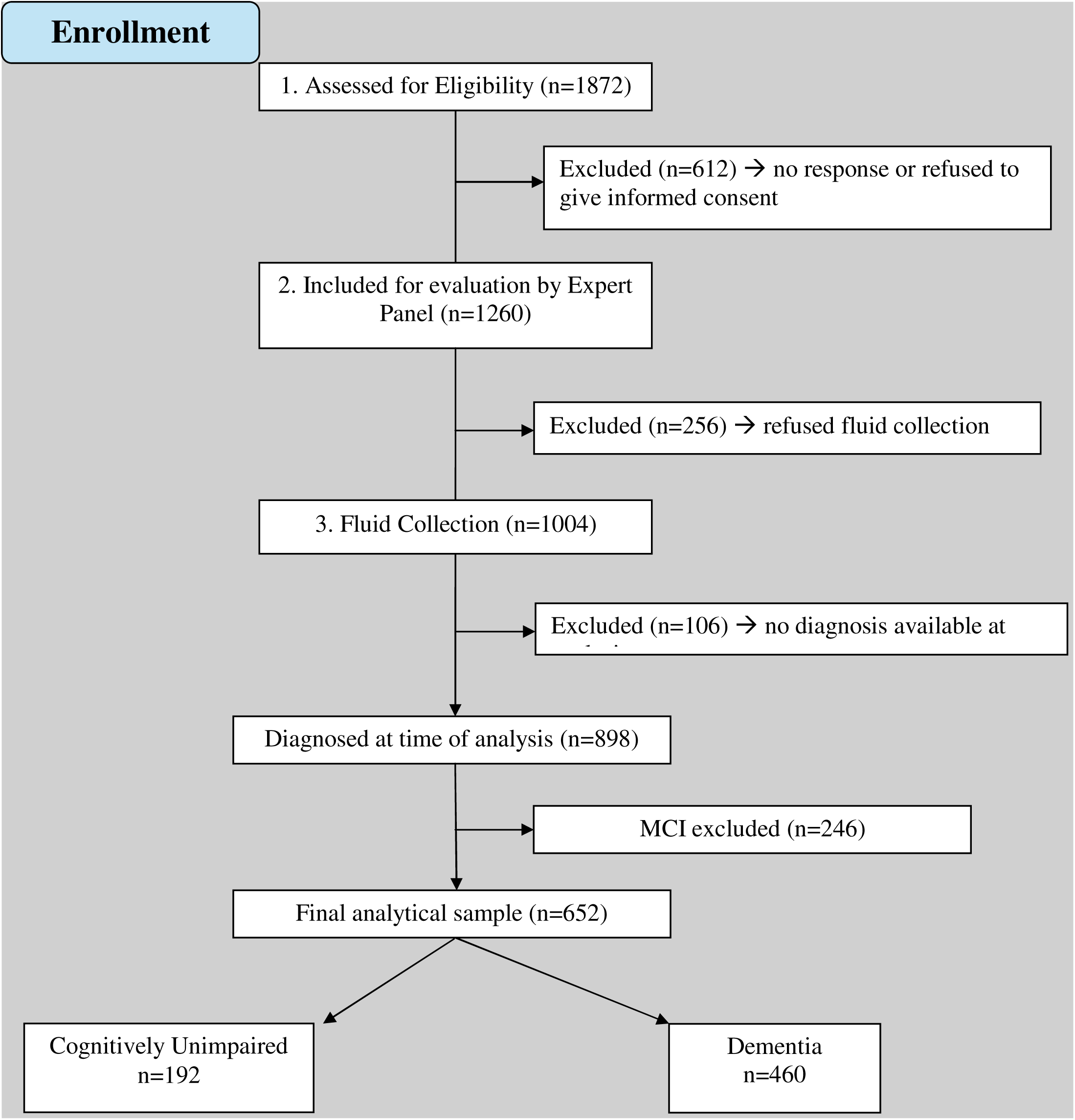
Participant flow diagram for the EVCD-RDC cohort and analytic sample selection. A total of 1872 individuals were assessed for eligibility. Of these, 612 were excluded because they did not respond or declined to provide informed consent, leaving 1260 participants evaluated by the expert diagnostic panel. Among these, 256 participants declined fluid collection, resulting in 1004 participants with completed biological sampling. At the time of analysis, 106 participants did not yet have a confirmed diagnosis and were excluded. Among the remaining 898 participants with available diagnoses, individuals classified as having mild cognitive impairment (MCI; n=246) were excluded because MCI represents an intermediate stage between normal cognition and dementia. The final analytic sample therefore included 652 participants classified as cognitively unimpaired or with dementia.

Predicted cognitive scores were derived using linear regression models including screening, functional, cognitive, and intelligence test scores. Outcome variables included scores from the screening, cognitive, intelligence, and functional measures; predictors included age, sex, and years of education. Regression specifications and cohort norms are described in our cohort profile.^17^ Of 1872 recruited participants, 612 declined consent. After adjudication of screening results, interviews, and neurological examinations, 1260 participants remained eligible. The final sample included 1004 participants. At the time of analysis, 898 participants had received a clinical diagnosis, while 106 participants had not yet completed diagnostic evaluation. Analyses were restricted to participants with available diagnoses (n=898). Participants diagnosed with mild cognitive impairment (MCI; n=246) were excluded because they represent an intermediate stage between normal cognition and dementia, leaving a final analytic sample of 652. The study was approved by the Ethics Committee/Institutional Review Board of the University of Kinshasa.

### Measures

The EVCD-RDC protocol included,^17^ the National Alzheimer’s Coordinating Center Uniform Data Set version 3 (UDS-3), dementia screening tools, the African Neuropsychology Battery–Short Version (ANB-SV), intelligence tests, and questionnaires addressing SSA-specific factors.

The UDS-3 comprises standardized forms capturing: participant demographics (A1), informant demographics (A2), family history (A3), medications (A4), health history (A5), physical examination (B1), Clinical Dementia Rating (CDR; B4), behavioral and neuropsychiatric symptoms (Neuropsychiatric Inventory Questionnaire, NPI-Q; B5), neurological examination (B8), clinician diagnosis (D1), and clinician-assessed medical conditions (D2)^32^. Dementia screening instruments included the Montreal Cognitive Assessment (MoCA), the CDR, and the IDEA functional scale, enabling harmonization with international cohorts.

The ANB-SV was developed to capture culturally relevant cognitive domains in SSA and included tasks of attention (Mancala Test), working memory (African Market Test), visuospatial perception (African Face Perception Test), learning and memory (African List Memory Test, African Story Memory Test, and a Visuospatial Memory Test), and executive functions (African Proverb Test and Card Game Test). Additional questionnaires assessed context-specific factors in SSA, including resilience, poverty, illiteracy, air pollution, and cardiovascular health. Composite indices for resilience and poverty were derived from questionnaire totals.

All instruments were culturally adapted and translated into French and Lingala using forward–backward translation, followed by pre-piloting and refinement before implementation. The use of UDS-3 and related instruments was intended to harmonize findings with previous dementia cohorts^17,18^ and align with international standards while maintaining local cultural validity.

#### Modifiable Risk Factor Measures

Guided by the 2024 Lancet Commission on dementia prevention, intervention, and care,^3^ we operationalized the following modifiable risk factors for dementia: educational attainment, smoking, excessive alcohol consumption, diabetes, hypertension, obesity, depression, social isolation, and air pollution. In addition to risk factors identified by the 2024 Lancet Commission, we included context-specific exposures (poverty, war experience, traumatic events, sleep disorders) given their high prevalence in the DRC. (Table 1).

**Table 1.**
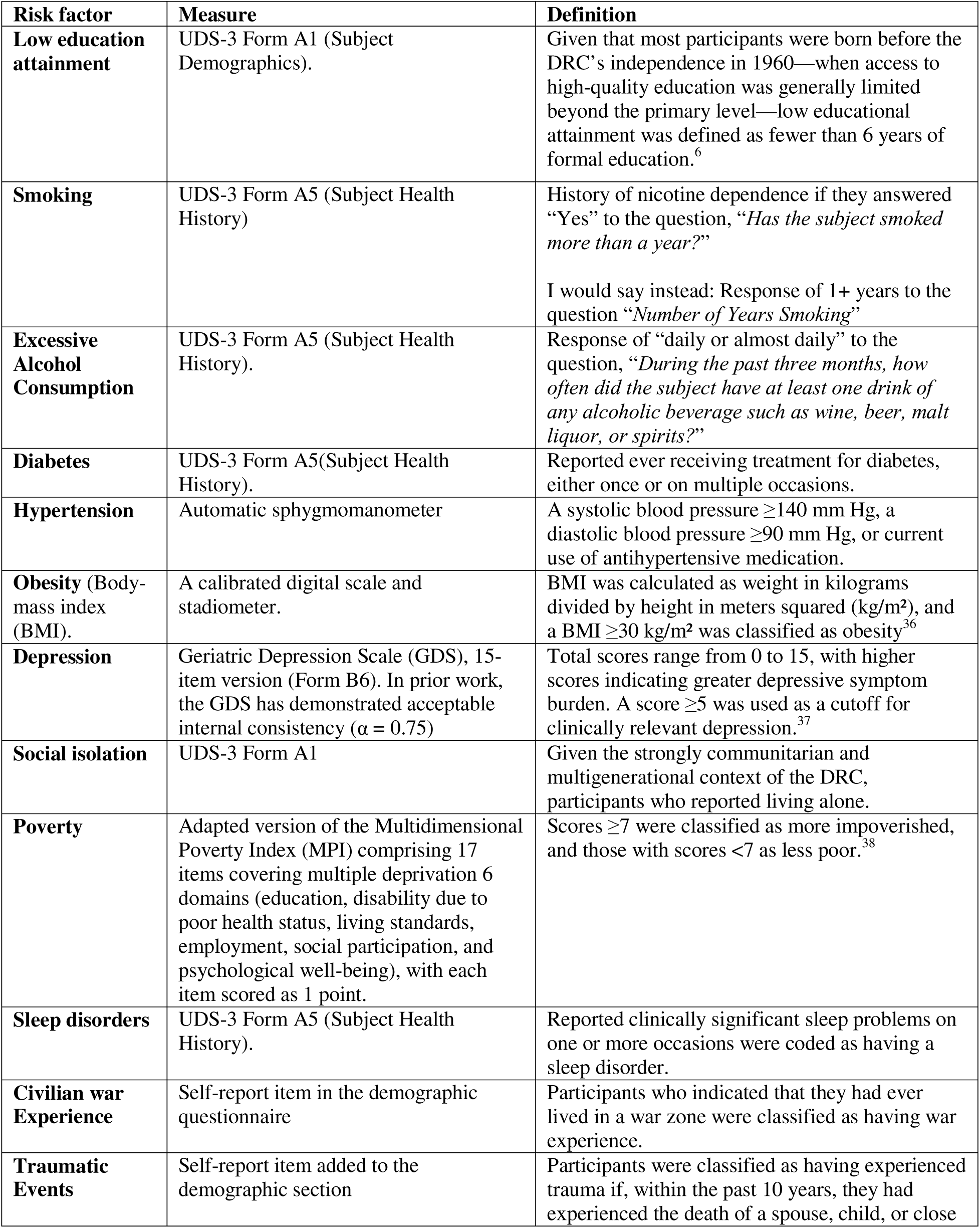
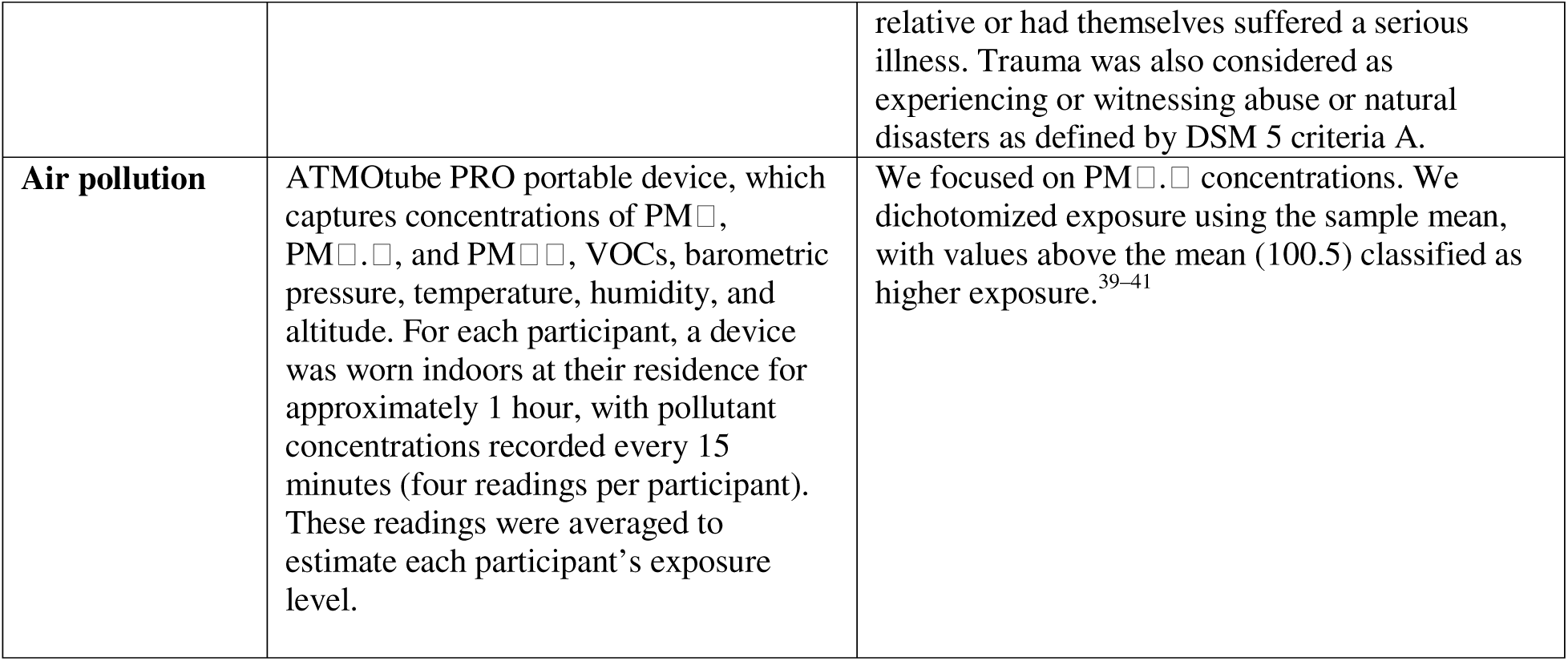
Measures and definitions of dementia risk factors.

### Data analysis

Analyses were conducted in R version 4.4.0. Descriptive statistics were generated for demographics (age, sex, and race), cognitive screening results, and modifiable risk factor variables. Categorical variables are presented as frequencies and percentages, and continuous variables (age and education) were summarized as medians with interquartile ranges (IQR). Variables with substantial missingness (hypertension, obesity, air pollution) were not imputed because missing values could not be assumed to indicate absence of exposure. Therefore, we used complete case analysis for odds ratio estimation. Logistic regression compared cognitively unimpaired and dementia groups. Unadjusted odds ratios for individual modifiable risk factors were calculated to show crude associations. Adjusted odds ratios (aORs) and 95% confidence intervals (CIs) were estimated controlling for age, sex, and education to account for potential confounding. Two-sided p values <0.05 were considered statistically significant.

PAFs were estimated for each risk factor using observed prevalence and measures of association. For a given risk factor with prevalence (P_e_) and relative risk estimate (RR) (approximated using odds ratios), the PAF was calculated as:

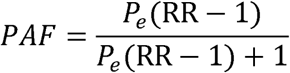

Unweighted, weighted, and combined PAFs were reported. Combined PAF estimates were calculated to reflect the overall attributable burden of dementia due to all risk factors noted. Weighted PAFs incorporated communality estimates (c) to account for overlap between correlated risk factors and avoid double counting of shared variance:

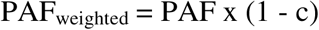

Potential impact fractions (PIFs) were then computed to estimate the expected proportional reduction in dementia burden under a hypothetical 15% proportional reduction in each risk factor prevalence. For baseline prevalence (P_e_) and reduced prevalence (P_e_*), PIF was calculated as:

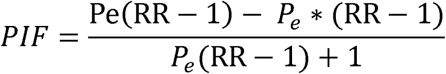

Projected reductions in dementia cases were extrapolated to the Democratic Republic of Congo population aged ≥65 years using an assumed dementia prevalence of 6.2% ^4^. The expected number of cases prevented was estimated as:

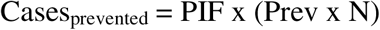

where (Prev) is dementia prevalence (6.2%) and (N) is the population aged ≥65 years in the DRC (2,702,000 individuals).

This study was reported in accordance with the STROBE reporting guidelines for observational studies. The funder had no role in study design, data collection, data analysis, data interpretation, or writing of the report. The corresponding author had full access to all data and had final responsibility for the decision to submit for publication.

## RESULTS

Demographic characteristics, cognitive screening results, and risk factor distributions by neurological status are shown in Table 1. The sample included 652 participants: 192 cognitively unimpaired (CU, 29%) and 460 with suspected dementia (71%). The high proportion of dementia cases reflects recruitment from settings enriched for cognitive concerns and does not represent population prevalence. Median age was 73.2 years (SD 6.1) and median education was 9.5 years (SD 4.9). Females represented 64% of the sample (n = 415). Participants with dementia were older and had fewer years of education than CU participants (p = 0.0001). Low education (p < 0.0001), depression (p = 0.020), social isolation, and poverty (p < 0.0001) were more common among participants with dementia.

**Table 2.** Characteristics of the Cohort Stratified by Dementia Status.

| **Variables,**<br>**n (%)** | **Overall**<br>**(n=652)** | **CU**<br>**(n=192; 29%)** | **Dementia**<br>**(n=460; 71%)** | **OR (95% CI)** | **p-value** |
| --- | --- | --- | --- | --- | --- |
| **Demographics** |  |  |  |  |  |
| Age, median (IQR) | 73.2 (6.1) | 71.1 (5.1) | 74.1 (6.2) | 1.1 (1.0, 1.1) | 0.0001 |
| Female | 415 (64%) | 112 (58%) | 303 (66%) | 0.74 (0.46, 1.2) | 0.20 |
| Education, median (IQR) years | 9.5 (4.9) | 11.8 (3.8) | 8.5 (4.9) | 0.84 (0.80, 0.89) | <.0001 |
| **Cognitive Screening** |  |  |  |  |  |
| MOCA | 20.2 (4.7) | 24.3 (2.4) | 17.8 (4.0) | 0.55 (0.46, 0.64) | <.0001 |
| CDR | 4.0 (3.8) | 1.7 (1.7) | 4.9 (4.0) | 1.5 (1.4, 1.7) | <.0001 |
| IDEA | 26.0 (6.6) | 28.9 (4.2) | 24.6 (7.1) | 0.90 (0.87, 0.94) | <.0001 |
| **Modifiable Risk Factors** |  |  |  |  |  |
| Low Education | 140 (22%) | 12 (6.3%) | 128 (28%) | 5.1 (2.6, 10.7) | <.0001 |
| Smoking | 42 (6.4%) | 18 (9.4%) | 24 (5.2%) | 0.67 (0.36, 1.4) | 0.29 |
| Alcohol Consumption | 13 (2.0%) | 5 (2.6%) | 8 (1.7%) | 1.2 (0.36, 4.0) | 0.82 |
| Diabetes | 56 (8.6%) | 12 (6.3%) | 44 (9.6%) | 1.9 (0.96, 4.1) | 0.074 |
| Sleep Disorder | 127 (20%) | 37 (19%) | 90 (20%) | 0.99 (0.62, 1.6) | 0.97 |
| Hypertension | 280 (43%) | 76 (40%) | 204 (44%) | 1.4 (0.96, 2.1) | 0.79 |
| Obesity* | 32 (4.9%) | 11 (5.7%) | 21 (4.6%) | 0.33 (0.11, 1.0) | 0.054 |
| Depression | 291 (45%) | 64 (33%) | 227 (49%) | 1.7 (1.1, 2.4) | 0.010 |
| Social Isolation | 32 (4.9%) | 15 (7.8%) | 17 (3.7%) | 0.41 (0.18, 0.91) | 0.027 |
| Poverty | 261 (40%) | 42 (22%) | 219 (48%) | 2.6 (1.7, 4.0) | <.0001 |
| War Experience | 270 (41%) | 74 (39%) | 196 (43%) | 1.3 (0.88, 1.9) | 0.21 |
| Traumatic Events | 381 (58%) | 104 (54%) | 277 (60%) | 1.4 (0.92, 2.0) | 0.12 |
| Air Pollution* | 45 (6.9%) | 11 (5.7%) | 34 (7.4%) | 1.1 (0.49, 2.6) | 0.82 |

\*Variables have a large number of missing data (obesity 78%, air pollution 56% missing). Individuals with missing data are not included because missingness cannot assume absence of risk.

Unadjusted odds ratios (ORs, 95% CI) and communalities are shown in Table 3. ORs ranged from 0.35 (0.14-0.88) for obesity to 5.9 (3.3-11.5) for low education. Communalities ranged from 65.1% (poverty) to 93.7% (obesity), indicating varying overlap between risk factors. (Table 3). Several exposures (smoking, obesity, social isolation) had ORs below 1.0, producing negative PAF estimates. This pattern may reflect residual confounding, survival bias, or limited statistical power.

**Table 3.** Odds Ratio of Modifiable Risk Factors and their communalities.

| **Risk Factors** | **Unadjusted OR**<br>**(95% CI)** | **Communality, %** |
| --- | --- | --- |
| Low Education | 5.9 (3.3, 11.5) | 78.0 |
| Smoking | 0.53 (0.28, 1.0) | 78.6 |
| Alcohol Consumption | 0.66 (0.22, 2.2) | 80.5 |
| Diabetes | 1.6 (0.85, 3.3) | 74.7 |
| Sleep Disorder | 1.0 (0.67, 1.6) | 83.0 |
| Hypertension | 1.2 (0.87, 1.7) | 86.9 |
| Obesity | 0.35 (0.14, 0.88) | 93.7 |
| Depression | 2.0 (1.4, 2.8) | 80.5 |
| Social Isolation | 0.45 (0.22, 0.94) | 87.5 |
| Poverty | 3.6 (2.4, 5.3) | 65.1 |
| War Experience | 1.2 (0.84, 1.7) | 70.7 |
| Traumatic Events | 1.3 (0.95, 1.9) | 67.8 |
| Air Pollution | 1.3 (0.64, 2.8) | 91.3 |

Table 4 shows prevalence and PAFs for each risk factor. Traumatic events were common (59.4%), whereas excessive alcohol consumption was rare. The largest weighted PAFs were poverty (18.4%, 95% CI 13.3–22.8), low education (11.3%, 7.3–15.3), depression (5.8%, 2.8–8.6), traumatic events (5.4%, 0.98–11.1), war exposure (2.1%, –2.1–6.4), diabetes (1.3%, 0.33–4.2), hypertension (1.1%, 0.81–3.1), and air pollution (0.39%, 0.52–1.9).Combined risk factors accounted for 37.3% (95% CI 14.3–55.6) of dementia cases. Poverty, low education, and depression contributed the largest fractions (Figure 2).

**Table 4.** Dementia risk factor prevalence and PAF: Prevalence (%) and PAF [%, (95% CI)

| **Risk Factors, %** | **Prevalence** | **Unweighted PAF** | **Weighted PAF** |
| --- | --- | --- | --- |
| Low Education | 21.8 | 51.6 (33.4, 69.6) | 11.3 (7.3, 15.3) |
| Smoking | 6.4 | -3.1 (-4.8, 0.11) | 0.67 (-1.0, 0.02) |
| Alcohol Consumption | 2.0 | 0.68 (-1.6, 2.4) | 0.13 (-0.31, 0.46) |
| Diabetes | 8.8 | 5.1 (-1.3, 16.6) | 1.3 (0.33, 4.2) |
| Sleep Disorder | 20.2 | 0.48 (-7.1, 10.5) | 0.08 (-1.2, 1.8) |
| Hypertension | 42.9 | 8.5 (-6.2, 23.6) | 1.1 (-0.81, 3.1) |
| Obesity | 22.7 | -17.2 (-24.1, -2.9) | -1.1 (-1.5, 0.18) |
| Depression | 44.8 | 29.9 (14.4, 44.4) | 5.8 (2.8, 8.6) |
| Social Isolation | 4.9 | -2.8 (-4.0, -0.31) | 0.35 (-0.5, -0.04) |
| Poverty | 43.7 | 52.7 (38.1, 65.3) | 18.4 (13.3, 22.8) |
| War Experience | 41.4 | 7.1 (-7.1, 21.8) | 2.1 (-2.1, 6.4) |
| Traumatic Events | 59.4 | 16.8 (-3.0, 34.5) | 5.4 (-0.98, 11.1) |
| Air Pollution | 15.6 | 4.4 (-6.0, 21.9) | -0.39 (-0.52, 1.9) |
| Combined Factors* | N/A | 87.2 (34.6, 98.7) | 37.3 (14.3, 55.6) |

\*Combined PAF estimate is weighted by risk factor communality and represented individual-level risk.
Note. Prevalence estimates reflect available-case data for each risk factor and therefore may differ from overall prevalence reported in Table 2.

**Figure 2.**
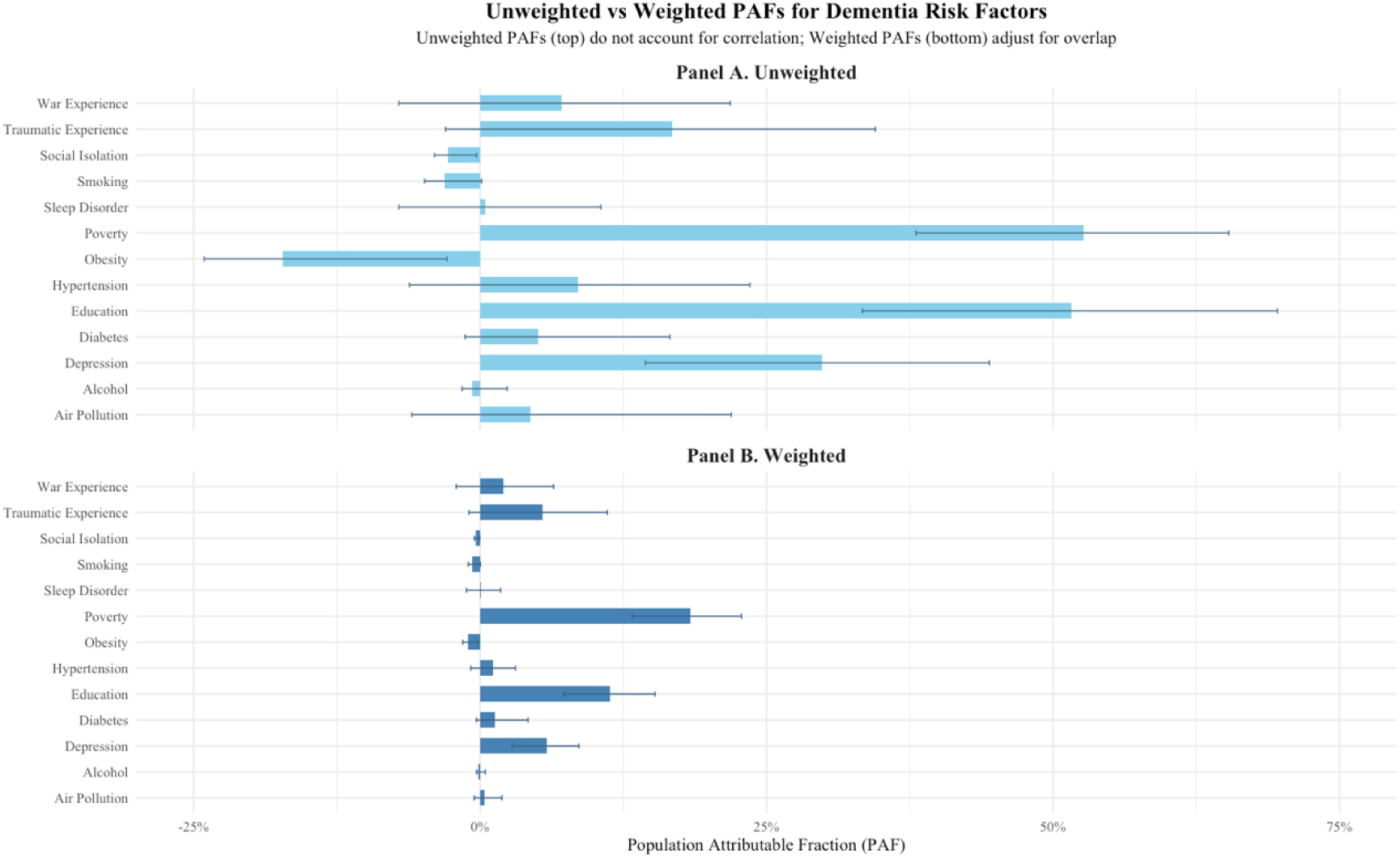
Panel A shows unweighted PAF estimates that do not account for correlations between risk factors. Panel B shows weighted PAF estimates adjusted for risk factor communality to account for overlap between exposures. Bars represent point estimates and error bars indicate 95% confidence intervals.

Table 5 shows potential impact fractions (PIFs) and estimated reductions in dementia cases with a 15% reduction in risk factors by 2025. Estimates assume a dementia prevalence of 6.2% and a projected population of 2,702,000 adults aged ≥65 years. T Reducing risk factors could lower dementia cases from 243,400 to 167,524. For example, a 15% reduction in poverty prevalence could reduce dementia prevalence by 2.8% (95% CI, 2.0–3.4%), corresponding to about 4600 cases (95% CI, 3300 –5700) in 2025. A 15% reduction in low education prevalence could be associated with a 1.7% (95% CI, 1.1–2.3%) lower dementia prevalence, corresponding to approximately 2800 cases (95% CI, 1800–3800). Similarly, a 15% reduction in depression prevalence could be associated with a 0.87% (95% CI, 0.42–1.3%) lower dementia prevalence, corresponding to about 1500 cases (95% CI, 700–2200). Overall, a 15% reduction in all risk factors combined could be associated with a 6.4% (95% CI, 2.1–10.8%) lower prevalence, corresponding to 10700 dementia cases (95% CI, 3600–18100) in 2025 (Table 5).

**Table 5.**
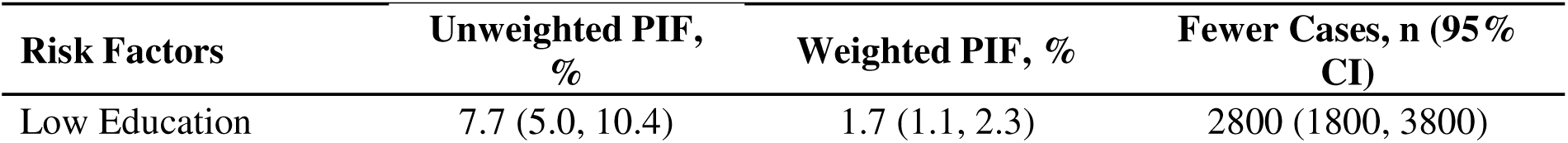
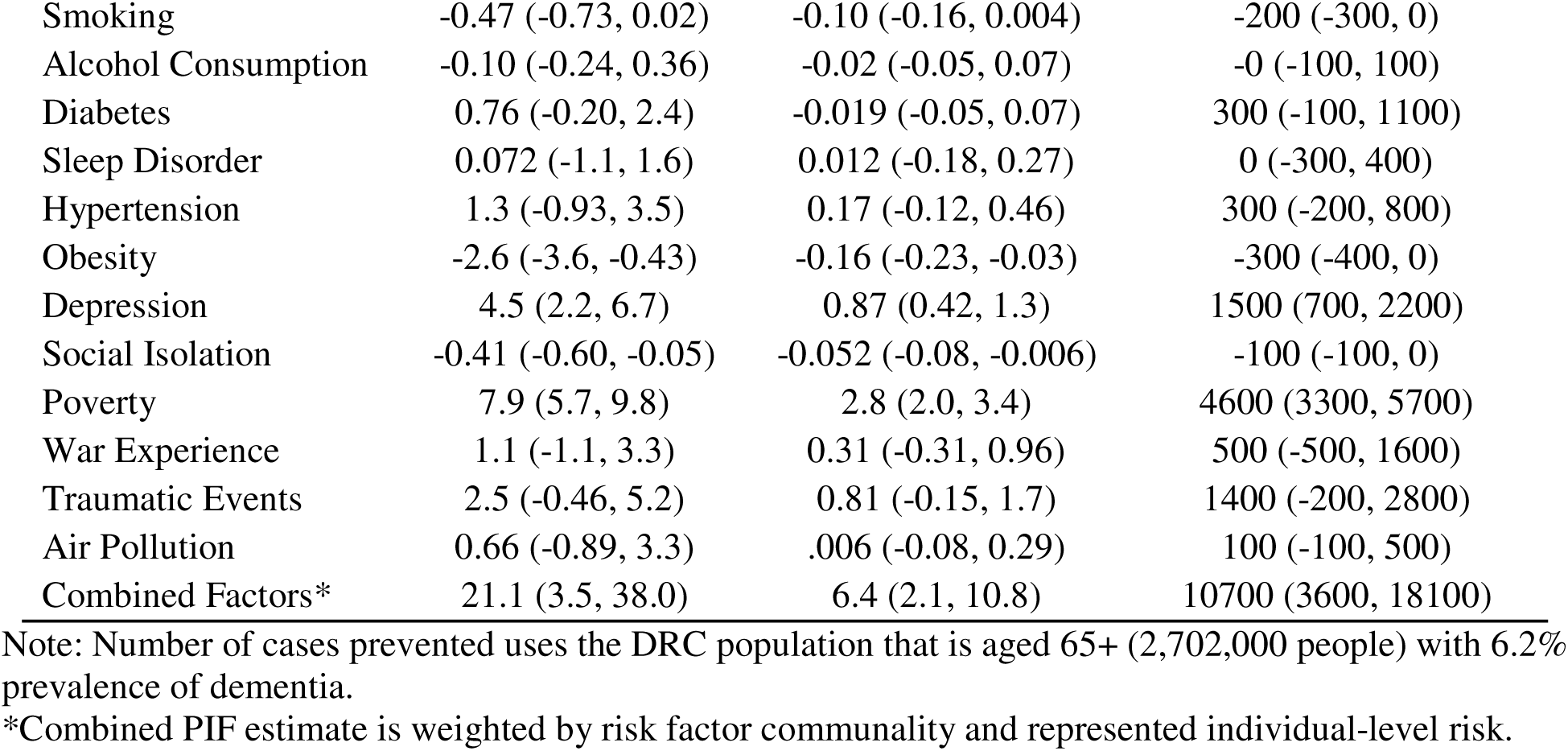
Potential impact fractions and estimated reduction in dementia cases in the Democratic Republic of the Congo associated with a 15% proportional reduction in risk factors by 2025.

| **Risk Factors** | **Unweighted PIF,**<br>**%** | **Weighted PIF, %** | **Fewer Cases, n (95%**<br>**CI)** |
| --- | --- | --- | --- |
| Low Education | 7.7 (5.0, 10.4) | 1.7 (1.1, 2.3) | 2800 (1800, 3800) |
| Smoking | -0.47 (-0.73, 0.02) | -0.10 (-0.16, 0.004) | -200 (-300, 0) |
| Alcohol Consumption | -0.10 (-0.24, 0.36) | -0.02 (-0.05, 0.07) | -0 (-100, 100) |
| Diabetes | 0.76 (-0.20, 2.4) | -0.019 (-0.05, 0.07) | 300 (-100, 1100) |
| Sleep Disorder | 0.072 (-1.1, 1.6) | 0.012 (-0.18, 0.27) | 0 (-300, 400) |
| Hypertension | 1.3 (-0.93, 3.5) | 0.17 (-0.12, 0.46) | 300 (-200, 800) |
| Obesity | -2.6 (-3.6, -0.43) | -0.16 (-0.23, -0.03) | -300 (-400, 0) |
| Depression | 4.5 (2.2, 6.7) | 0.87 (0.42, 1.3) | 1500 (700, 2200) |
| Social Isolation | -0.41 (-0.60, -0.05) | -0.052 (-0.08, -0.006) | -100 (-100, 0) |
| Poverty | 7.9 (5.7, 9.8) | 2.8 (2.0, 3.4) | 4600 (3300, 5700) |
| War Experience | 1.1 (-1.1, 3.3) | 0.31 (-0.31, 0.96) | 500 (-500, 1600) |
| Traumatic Events | 2.5 (-0.46, 5.2) | 0.81 (-0.15, 1.7) | 1400 (-200, 2800) |
| Air Pollution | 0.66 (-0.89, 3.3) | .006 (-0.08, 0.29) | 100 (-100, 500) |
| Combined Factors* | 21.1 (3.5, 38.0) | 6.4 (2.1, 10.8) | 10700 (3600, 18100) |

Note: Number of cases prevented uses the DRC population that is aged 65+ (2,702,000 people) with 6.2% prevalence of dementia.
\*Combined PIF estimate is weighted by risk factor communality and represented individual-level risk.

## DISCUSSION

In this community-based cross-sectional analysis of older adults in Kinshasa, approximately one-third of dementia cases were attributable to modifiable risk factors, primarily poverty and low educational attainment. Our analysis considered the recent Lancet Commission report^3^ and socioeconomic factors specific to the DRC. We estimated that 37.3% of dementia cases in the DRC may be attributable to modifiable risk factors. This estimate is consistent with the Lancet Commission, which reported that up to 40% of dementia cases globally could be prevented or delayed by modifying risk factors across the life course.^3,4^ However, the dominant risk factors identified differed from those emphasized in the Lancet Commission report. Using our previous study on dementia prevalence in the DRC^4^ and population projections for adults aged ≥65 years, we estimate that a 15% reduction in risk factor prevalence could correspond to approximately 10 700 fewer dementia cases. These findings highlight the growing dementia burden in low- and middle-income countries (LMICs), particularly in SSA. By 2050, an estimated 60–70% of people living with dementia worldwide will reside in LMICs.

The most prevalent modifiable risk factors were socioeconomic (poverty, low education, traumatic events, war exposure), metabolic (diabetes, hypertension), and environmental (air pollution). In contrast, midlife hypertension, obesity, and physical inactivity are among the primary modifiable risk factors identified in the United States.^33^ While many studies identify obesity as a dementia risk factor, our study found late-life obesity protective.^34^ Prior work suggests that late-life obesity may reflect nutritional reserves in contexts of frailty and poverty.^34^ Whereas metabolic factors dominate in high-income countries, socioeconomic factors appear more important in SSA. Our findings align with a study from Soweto, South Africa, showing higher rates of multidimensional poverty among adults with dementia than those without. Individuals classified as multidimensionally poor had more than twice the risk of dementia.^17^ Multidimensional poverty remains widespread in SSA.^18^ MPI and extreme poverty—defined by the World Bank Group (WBG) as living on less than $2.15 per day (purchasing power)—represent one of the most important characteristics of LMICs, ^6^ and of the DRC, where 73.5% of residents meet that definition.^7^ These findings underscore the importance of poverty as a key modifiable determinant of dementia risk in LMICs.

Comparisons with other SSA countries show similarities and differences in risk factor profiles.^8^ Low education was a stronger contributor in Mozambique, whereas poverty contributed the largest fraction in the DRC.In Mozambique, smoking was the second strongest risk factor (PAF 11.2%), followed by depression (PAF 6.5%) and hypertension (PAF 6.1%). In the DRC, the second strongest risk factor was low education (PAF 11.3%), followed by depression (PAF 5.8%) and traumatic events (PAF 5.4%). Metabolic risk factors (hypertension and diabetes) were also important in both countries. The PAF for diabetes was the same in both (1.3%), while the PAF for hypertension was higher in Mozambique (6.1%) compared to the DRC (1.3%). Obesity contributed a PAF of 0.9% in Mozambique but was negative in the DRC (–1.1%). These findings confirm the rising burden of non-communicable diseases in LMICs, reflecting an epidemiological transition from infectious and environmental risks to metabolic conditions. Over time, these metabolic factors are likely to become the leading dementia risk factors, as observed in Western, Educated, Industrialized, Rich, and Developed (WEIRD) countries.^35^

### Strengths and limitations

This study has several strengths. It is among the first analyses of PAFs for dementia risk factors in sub-Saharan Africa and incorporates both conventional cardiometabolic risk factors and structural determinants such as poverty, conflict exposure, and traumatic events. The study draws on a community-based cohort of older adults in Kinshasa and applies communality weighting to account for correlations when estimating combined PAFs. These findings may inform policy initiatives aimed at reducing dementia risk in the DRC. Such initiatives could target reductions in key modifiable risk factors.^33^

Several limitations should be considered when interpreting these findings. First, the cross-sectional design precludes causal inference, and reverse causation cannot be excluded for exposures such as depression, social isolation, or poverty. Second, recruitment was community-based but not probability-based, which may limit representativeness and introduce selection bias. Third, several exposures—including hypertension, obesity, and air pollution—had missing data, and complete-case analyses may have introduced bias if missingness was not random. Fourth, PAF estimates were derived using odds ratios from logistic regression; although dementia prevalence in the underlying population is relatively low, odds ratio–based PAF estimates may overestimate attributable risk. Levin’s formula also assumes independence of exposures, and although communality adjustments were applied, residual confounding and overlap between risk factors may remain. Finally, air pollution exposure was measured using short-duration indoor PM. sampling, which may not reflect long-term ambient exposure. These limitations should be considered when interpreting the magnitude and policy implications of these estimates.^31–33^

Despite these limitations, we demonstrated statistically how reducing modifiable risk factors could decrease dementia cases. However, concrete and context-specific interventions are needed for each risk factor in the DRC. For example, while the Finnish Geriatric Intervention Study to Prevent Cognitive Impairment and Disability (FINGER) emphasized diet, physical activity, cognitive training, and vascular risk management, interventions in Europe and USA—and particularly in the DRC—must prioritize reducing poverty, depression, traumatic events, and war-related stressors.

Future studies should include larger, representative samples across multiple regions of the DRC. They should rely on representative data, consider multiple exposures, and incorporate multivariate models and sensitivity analyses to adjust for confounding. More accurate national data on exposure prevalence and risk estimates are needed to strengthen the evidence base for dementia prevention strategies in LMICs.

### Conclusions

In conclusion, modifiable risk factors account for a substantial proportion of dementia cases in this population, with structural determinants such as poverty and education contributing the largest fractions. These findings underscore the importance of integrating socioeconomic and public health interventions into dementia prevention strategies in the DRC and other LMICs.

## RESEARCH IN CONTEXT

### Evidence before this study

Population attributable fraction analyses have been widely used to estimate the proportion of dementia cases that could be prevented through modification of known risk factors. The Lancet Commission on dementia prevention, intervention, and care estimated that up to 40% of dementia cases worldwide may be attributable to modifiable risk factors across the life course. However, most of the evidence informing these estimates originates from high-income countries. Data from low- and middle-income countries, particularly in sub-Saharan Africa, remain limited despite the region’s rapidly growing older population and increasing dementia burden. Furthermore, most prior analyses focus primarily on clinical and lifestyle risk factors and rarely incorporate broader social or structural determinants of health that may substantially influence dementia risk in low-resource settings.

### Added value of this study

This study provides one of the first estimates of population attributable fractions for dementia risk factors in the Democratic Republic of Congo. In addition to traditional vascular and lifestyle risk factors, our analysis incorporates structural and contextual determinants—including poverty, low educational attainment, exposure to traumatic events, and war exposure—that are highly prevalent in this setting but rarely included in dementia prevention models. Our findings suggest that approximately 37% of dementia cases in this population may be attributable to potentially modifiable risk factors, with poverty, low education, and depression contributing the largest fractions.

### Implications of all available evidence

These findings suggest that dementia prevention strategies in sub-Saharan Africa may require broader public health approaches that address social and structural determinants of health alongside conventional clinical risk factors. Policies aimed at poverty reduction, improving educational access, and strengthening mental health services may have substantial downstream effects on cognitive health in later life. Expanding dementia prevention frameworks to incorporate structural determinants may therefore be critical for effective population-level dementia prevention in low- and middle-income countries.

## Data Availability

All data produced in the present study are available upon reasonable request to the authors.

## CONTRIBUTORS

JI: Conceptualization, Data curation, Funding acquisition, Investigation, Methodology, Project administration, Resources, Software, Supervision, Validation, Visualization, Writing – original draft, Writing – review & editing. SP: Writing – original draft, Writing – review & editing. MS: Formal analysis, writing – original draft, writing – review & editing. CO: Writing – review & editing. EE: Writing – review & editing. GG: Writing – review & editing. NT: Writing – review & editing. IK: Writing – review & editing. SM: Writing – review & editing. LM: Writing – review & editing. AS: Writing – review & editing. JR: Writing – review & editing. BC: Writing – review & editing. AL: Writing – review & editing. JK: Writing – review & editing. AB: Writing – review & editing. AJ: Writing – review & editing. AG: Writing – review & editing. AA: Writing – review & editing. JI and MS directly accessed and verified the underlying data reported in the manuscript. All authors had access to the data, reviewed the manuscript, and approved the final version for submission.

## DATA SHARING STATEMENT

Deidentified participant data used in this analysis will be made available upon reasonable request to the corresponding author beginning at the time of publication. Supporting documents, including the study protocol and statistical analysis plan, will also be available upon request. Data access will require approval from the study investigators and a signed data use agreement.

## ACKNOWLEDGEMENTS

This work was supported by the National Center for Advancing Translational Sciences of the National Institutes of Health under Award Number UL1TR002378. The funding source had no role in the study design, data collection, data analysis, data interpretation, writing of the report, or the decision to submit the manuscript for publication.

We thank the study participants and community collaborators who contributed to data collection in Kinshasa, Democratic Republic of Congo. We are also grateful to the Centre Médical de Kinshasa (CMK), which provided the necessary infrastructure for data collection and storage.

